# SARS-CoV-2 N gene dropout and N gene Ct value shift as indicator for the presence of B.1.1.7 lineage in a widely used commercial multiplex PCR assay

**DOI:** 10.1101/2021.03.23.21254171

**Authors:** P Wollschlaeger, N Gerlitz, D Todt, S Pfaender, T Bollinger, A Sing, A Dangel, N Ackermann, K Korn, A Ensser, E Steinmann, M Buhl, J Steinmann

**Author notes:** **Correspondence to:** Joerg Steinmann, MD, Institute of Clinical Hygiene, Medical Microbiology and Infectiology, Paracelsus Medical University, Prof.-Ernst-Nathan-Str. 1, 90419 Nuremberg, Germany. P. Wollschläger, N. Gerlitz and D. Todt contributed equally to the work. These authors share senior authorship (M Buhl, J Steinmann).

## Abstract

**Objectives:** Increased importance in detection and surveillance of SARS-CoV-2 has been demonstrated due to the emergence of variants of concern (VOCs). In this study we evaluated if a commercially available real-time SARS-CoV-2 PCR assay can identify B.1.1.7 lineage samples by a specific N gene dropout or Ct value shift compared to the S or RdRP gene.

**Methods:** Patients samples with confirmed B.1.1.7 variant by whole-genome sequencing and variant-specific PCR (n=48) and non-B.1.1.7 samples (n=53) were tested by the Allplex™ SARS-CoV-2/FluA/FluB/RSV™ PCR assay for presence of S, RdRP and N gene of SARS CoV-2. The N gene coding sequence of SARS-CoV-2 with and without D3L mutation (specific for B.1.1.7) were cloned into pCR®-TOPO vectors and Allplex™ SARS-CoV-2/FluA/FluB/RSV™ PCR assay was performed.

**Results:** All studied B.1.1.7 patient samples showed significantly higher Ct values (Δ 6-10, N-gene dropout on Ct values >29) in the N gene compared to the respective values of S and RdRP gene. Receiver operating characteristic (ROC) curve analysis resulted in 100% sensitivity and specificity for ΔCt N/RdRP and ΔCt N/S. As a result of the reversed genetic experiments we found also the shift in Ct values for the 3L variant N-gene.

**Conclusions:** N gene dropout or Ct value shift is specific for B.1.1.7 positive samples using the Allplex™ SARS-CoV-2/FluA/FluB/RSV PCR assay. This approach can be used as a rapid tool for B.1.1.7 detection in single assay high throughput diagnostics.

## Introduction

Severe acute respiratory syndrome coronavirus-2 (SARS-CoV-2), the cause of the life-threating COVID-19 disease, has rapidly spread rapidly worldwide since December 2019. This positive-sense single-stranded RNA virus, a member of the beta-coronavirus subfamily. Since transmission to humans the virus undergoes adaptive evolution, resulting in genetic variation that can be a challenge particularly in molecular diagnostics, which are based on the detection of small regions of the viral genome. To this date, several major variants of concern (VOCs) of SARS-CoV-2 have been reported. Currently, (i) B.1.351 (first detected in South Africa) [1], (ii) P.1, (Brazil) [2] and (iii) B.1.1.7 (United Kingdom) [3] are among the most important ones. The B.1.1.7 variant has been associated with an increased mortality [4] and risk of transmission [5] and is rapidly expanding its geographic range worldwide; therefore, early identification of this variant in patients may help reduce further spread of infection. Each lineage is characterized by a combination of single mutations across the SARS-CoV-2 genome, and while some are shared between the aforementioned lineages, others are specific for one individual lineage. For example, amino acid exchange N501Y in the SARS-CoV-2 spike (S) protein is present in all three mentioned lineages, whereas the additional deletion H69/V70 in the same protein is present in B1.1.7, but not in the other two VOCs B.1.351 and P.1 [3]. The B.1.1.7 has also a specific mutation in the N gene (D3L) which can be targeted by PCR assays.

Multiplex PCR is considered as gold standard for SARS-CoV-2 detection, and dual or triple target approaches are recommended. However, identification of viral variants is not intended with regular PCR kits. The presence of SARS-CoV-2 variants in a patient sample can potentially change the performance of the SARS-CoV-2 assay. Occasional dropouts of various genes included as targets in SARS-CoV-2 multiplex PCR approaches have been reported, affecting the S gene [6,7], N gene [8,9] or E gene [10], which may allow presumptive identification of specific lineages. While rapid spread of VOC lineages is an imminent danger in Europe and worldwide, rapid detection of VOCs is limited by the turnaround time and costs or availability of methods such as next-generation sequencing (NGS) or variant specific PCR, respectively. Here, we report an approach which allows for presumptive identification of the B.1.1.17 lineage by calculating a score from the Ct values of the target genes included in the Allplex™ SARS-CoV-2/FluA/FluB/RSV Assay (Seegene, South Korea), thus greatly speeding up the time to result of laboratory diagnosis of this lineage.

## Method and Results

Using a real-time (RT) PCR assay with SARS-CoV-2 detection based on a triple target approach (S gene, RdRP gene, and N gene), we found N gene-specific changes in early 2021 in a large clinical diagnostics laboratory in Southern Germany. A total of 48 consecutive samples all showing N gene dropouts (n=6) or N gene cycle threshold (Ct) value shifts (n=42) was collected for further analysis aiming at lineage-specific assignment of the respective SARS-CoV-2 strains. To this end, either whole-genome sequencing (n=23) (Illumina MiSeq and Novaseq 6000) or variant specific PCR (n=25) (Novaplex™ SARS-CoV-2 Variants I Assay, Seegene) were performed, confirming all samples with a Δ N/RdRP or N/S score > 5 as belonging to the B.1.1.7 lineage. In addition, a selection of 58 samples with unsuspicious Δ N/RdRP or N/S scores (< 5) sampled during the same period of time was also subjected to lineage-specific analysis (Novaplex™ SARS-CoV-2 Variants I Assay), confirmed as non-B.1.1.7 lineages (B.1.1.317 (n=2), B.1.351 (n=1), N501Y+, E484K+ (n=1), N501Y-, E484K+ (n=1) and N501Y-, Del. H69/V70- (n=53), reflecting the local epidemiology in early 2021 (Supplemental Table 2).

The observed N gene Ct shift or N gene dropout was detectable regardless if samples were strongly or weakly positive for SARS-CoV-2, as evident from the broad range of Ct values for B.1.1.7 and non-B.1.1.7 (Figure 1A). Receiver operating characteristic (ROC) curve analysis resulted in 100% sensitivity and specificity for ΔCt N/RdRp and ΔCt N/S (95% confidence interval (CI) sensitivity 91.62%-100% and selectivity 93.79%-100), while there was no such effect for ΔCt S/RdRP genes, also reflected by differences in areas under the curve (AUC) (Figure 1B). In addition, the Ct value shifts observed for both N/RdRP and N/S genes allows for meaningful classification of B.1.1.17 versus non-B.1.1.17 (no overlapping/outliers). This is also supported be the homogeneity for N/RdRP and N/S gene Ct values (visualized in linear regression analysis in Figure 1 C).

**Figure 1:**
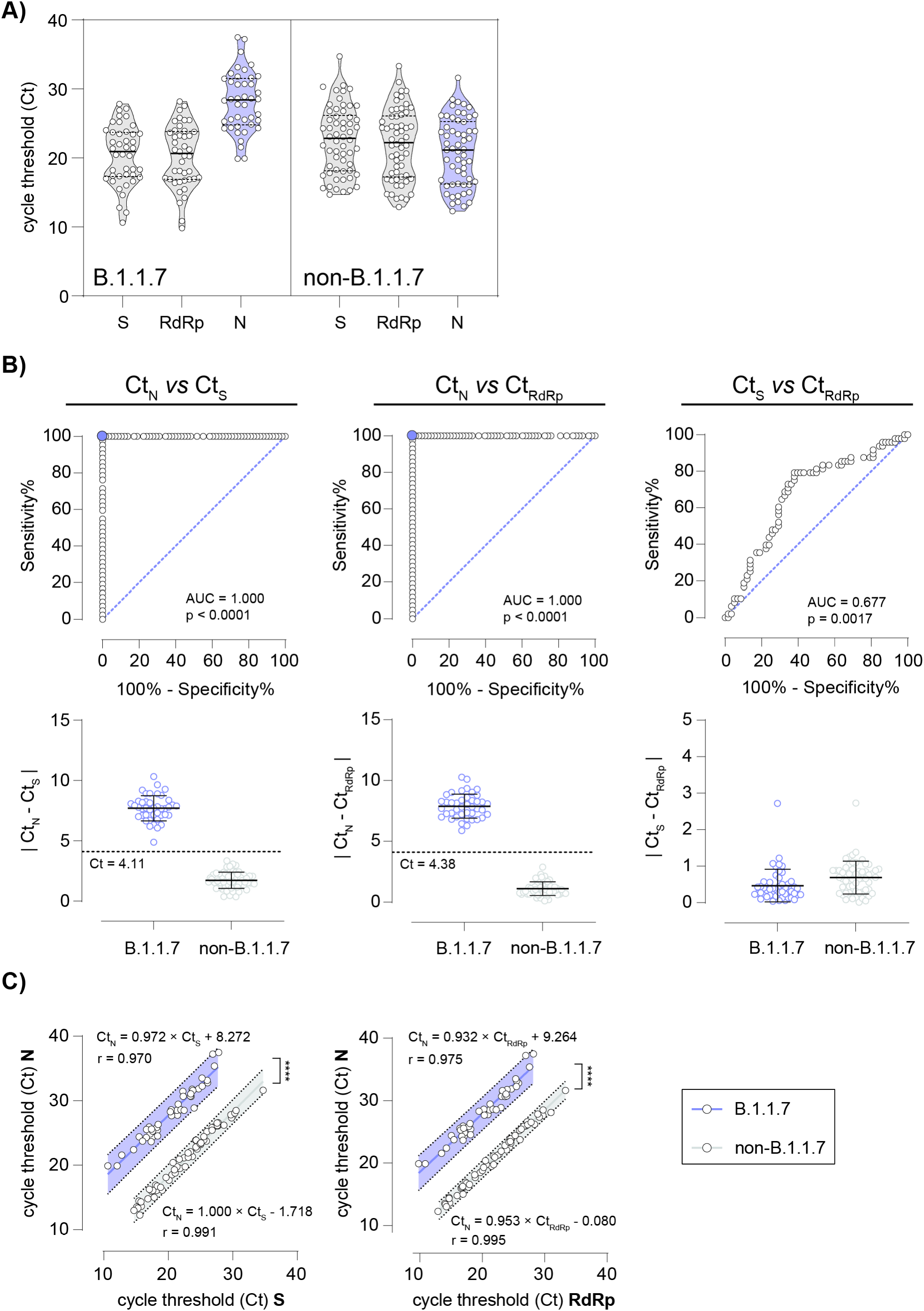
The Allplex™ SARS-CoV-2/FluA/FluB/RSV Assay reliably discriminates SARS-CoV-2 B.1.1.7 from non-B.1.1.7 positive patient specimens. **A)** Violin plots with median (solid black lines), quartiles (dashed black lines) and individual data points (open circles) of determined cycle thresholds (Ct) values for B.1.1.7 and non-B.1.1.7 positive patient specimens in RT-PCR for three genes, S, RdRp and N. **B)** Receiver operating characteristic (ROC) curves discriminating absolute differences in Ct values of B.1.1.7 and non-B.1.1.7 positive patient specimens between N gene and S gene (left panel), N gene and RdRp gene (middle panel) and S gene and RdRp gene (right panel). Lower respective panels display differences in Ct values; dashed lines indicate Ct with 100% sensitivity and specificity as determined in ROC curves. **C)** Linear regression of Ct values of N gene and S gene (left panel) and N gene and RdRp gene of B.1.1.7 (blue line and 99% prediction band) and non-B.1.1.7 (gray line and 99% prediction band) positive patient specimens, respectively. No significant difference in slope is determined, y-intercepts (elevations) differ significantly (**** p < 0.0001).

In order to assess the specificity of our findings, a subset of the samples with N gene Ct shift or N gene dropout were tested by two other commercially available SARS-CoV-2 PCR assays. Interestingly, neither the Allplex™ 2019-nCoV (Seegene) nor the VIASURE SARS-CoV-2 (CerTest, for BD MAX™ System) showed N gene Ct shift or N gene dropout.

To elucidate the underlying molecular mechanism resulting in the observed N gene dropout or Ct value shift, we cloned the N gene coding sequence with and without D3L mutation into pCR®-TOPO vectors. Next, each plasmid was diluted to an initial copy number of 125*10^8 afterwards diluted to 1:10, 1:100 and 1:1000 dilutions. To imitate extraction and test conditions, 0.5 µl of each dilution was then mixed with 500 µl viral transport medium containing a negative tested nasopharyngeal swab. Nucleic acid extraction was performed on Seegene Nimbus IVD using the STARMag 96 × 4 Viral DNA/RNA 200 C Kit (Seegene). Amplification and detection of SARS-CoV-2 was performed with Allplex™ SARS-CoV-2/FluA/FluB/RSV. For variant specific PCR, Novaplex™ SARS-CoV-2 Variants I assay was performed.

Again, we found a shift in Ct values for the 3L variant N-gene using the Allplex™ SARS-CoV-2/FluA/FluB/RSV assay. This confirmed that the D3L mutation can be discriminated by the Allplex™ SARS-CoV-2/FluA/FluB/RSV assay, but not by the Allplex™ 2019-nCoV test (Figure 2A, B).

**Figure 2:**
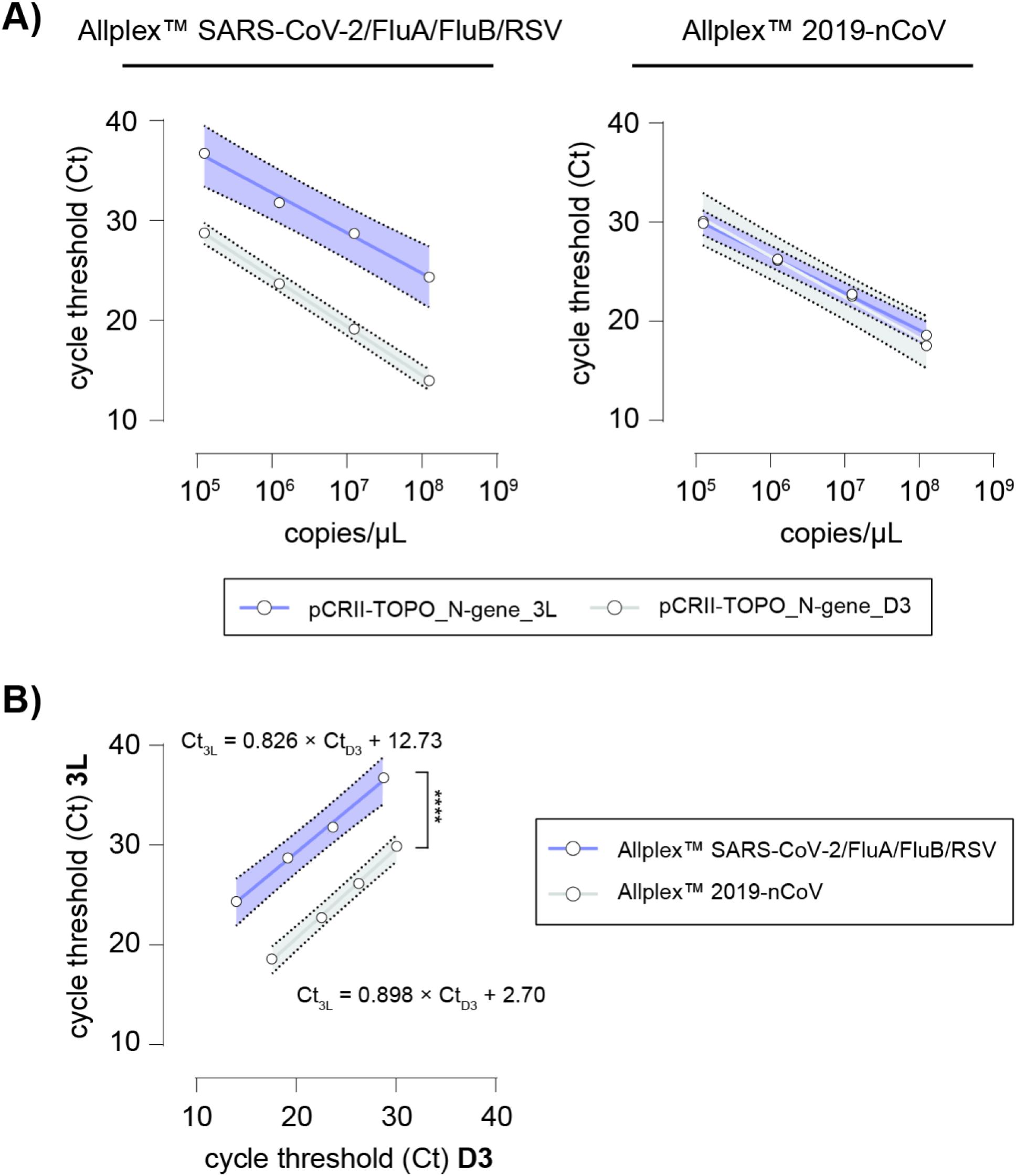
The Allplex™ SARS-CoV-2/FluA/FluB/RSV Assay discriminates SARS-CoV-2 N gene 3L from D3 plasmids, while the Allplex™ 2019-nCoV Assay does not. **A)** Standard curves of pCRII-TOPO_N-gene_3L (blue line and 99% prediction band) and _D3 (gray line and 99% prediction band) plasmid titrations using the Allplex™ SARS-CoV-2/FluA/FluB/RSV assay (left panel) and the Allplex™ 2019-nCoV assay (right panel), respectively. **B)** Linear regression of Ct values of pCRII-TOPO_N-gene_3L and _D3 assessed using the Allplex™ SARS-CoV-2/FluA/FluB/RSV assay (blue line and 99% prediction band) and the Allplex™ 2019-nCoV assay (gray line and 99% prediction band). No significant difference in slope is determined, y-intercepts (elevations) differ significantly (**** p < 0.0001).

## Discussion

### N gene dropout or Ct value shift are specific for B.1.1.7 positive samples

We were able to demonstrate that all confirmed B.1.1.7 samples have an N gene dropout or Ct value shift compared to the S or RdRP gene for SARS-CoV-2 by the Allplex™ SARS-CoV-2/FluA/FluB/RSV assay. These N gene dropouts or Ct value shifts were caused by D3L mutation in the N gene of SARS-CoV-2 which was confirmed by reverse genetics. In contrast, strains of other lineages were not affected by N gene dropouts or Ct value shifts. Mutations in the N gene are known to occur less often than in other regions of the SARS-CoV-2 genome [11–15]. To the best of our knowledge, this is the first report of N gene dropout or Ct value shift with diagnostic relevance as a marker of the B.1.1.7 lineage (Allplex™ SARS-CoV-2/FluA/FluB/RSV Assay), similar to what has previously been reported for S gene dropouts for TaqPath COVID-19 assay (ThermoFisher, US). However, the N gene dropout has a higher specificity for the B.1.1.7 lineage, as it does not occur in N501Y-, ds69-70 strains (e.g. lineages from mink and others).

### Allplex™ SARS-CoV-2/FluA/FluB/RSV Assay can be used as fast detection tool for presumptive identification of the B.1.1.7 lineage

Detection of SARS-CoV-2 variants has become an additional task for many laboratories, which can be a time critical challenge. As whole-genome sequencing is the gold standard to detect, track and monitor virus variants (surveillance), this method is neither accessible everywhere nor easy-to-perform. Especially in clinical contexts, fast identification of VOC lineages is crucial for patient management in both inpatient and outpatient settings (infection control and prevention measures including contact tracing). While most laboratories have meanwhile implemented workflows to deliver SARS-CoV-2 RT-PCR results in a timely manner, variant specific discrimination of positive samples requires additional resources and thus delays reporting of VOCs. This counteracts public health strategies particularly aiming at slowing down the spread of VOCs because of their increased transmissibility.

In addition to the detection of four viral respiratory pathogens (SARS-CoV-2, Influenza A and B plus RSV), the Allplex™ SARS-CoV-2/FluA/FluB/RSV Assay allows for presumptive assignment of SARS-CoV-2 positive samples to the B.1.1.7 lineage in a one-step approach by using the proposed Δ N/RdRP or N/S score. However, confirmation of this presumptive result is warranted as the Allplex™ SARS-CoV-2/FluA/FluB/RSV Assay is not intended for detection of VOCs by the manufacturer. Furthermore, the proposed Δ N/RdRP or N/S score cannot be applied for detection of VOCs other than the B.1.1.7 lineage.

In conclusion, this approach can be used as a rapid tool for B.1.1.7 detection and surveillance in high throughput diagnostics.

## Data Availability

The authors confirm that the data supporting the findings of this study are available within the article its supplementary materials.

## Institutional Review Board Statement

The study was conducted according to the guidelines of the Declaration of Helsinki and approved by the Institutional Review Board, Paracelsus Medical University, Nuremberg,Germany (reference number IRB-2021-008; 22 March 2021).

## Authors’ contributions

Designed experiments: WP, GN, TD, PS, SE, SJ. Performed experiments: WP, GN, TD. Provided important samples and data: WP, GN, TD, PS, BT, SA, DA, AN, KK, EA, SE, BM, SJ. Wrote the manuscript: WP, GN, TD, BM, SJ.

## Acknowledgment

We would like to thank all members of the Institute of Clinical Hygiene, Medical Microbiology and Infectiology, Paracelsus Medical University for support and fruitful discussion.

---

**Supplemental data 1A, B, C, D. A)**. Linear regression of Ct values of S gene and RdRp gene of B.1.1.7 (left panel, blue line and 99% prediction band) and non-B.1.1.7 (right panel, gray line and 99% prediction band) positive patient specimens. **B and C**). Included samples (n=106) with Ct values of the S, RdRP and N gene by the Allplex™ SARS-CoV-2/FluA/FluB/RSV assay with the corresponding results of whole genome sequencing or variant specific PCR. N501Y+, Del. H69/V70+ indicate presence of lineage B.1.1.7 **D.)** Nucleotide sequence alignment (MUSCLE) of N gene sequences from two exemplary patient samples (non-B.1.1.7 and B.1.1.7, respectively) and pCRII-TOPO_N-gene_D3 and _3L plasmids. Only the region containing the D3L mutation is shown (nucleotide position 1-12), with D3 highlighted in green (non-B.1.1.7) and 3L (B.1.1.7) highlighted in red. Translated amino acid sequences are shown above and below the nucleotide sequences. N gene sequences from the patient samples were extracted from whole-genome sequences.

