## Supplemental Data for "SARS-CoV-2 N gene dropout and N gene Ct value shift as indicator for the presence of B.1.1.7 lineage in a widely used commercial multiplex PCR assay"

**Supplemental data 1A, B, C, D. A).** Linear regression of Ct values of S gene and RdRp gene of B.1.1.7 (left panel, blue line and 99% prediction band) and non-B.1.1.7 (right panel, gray line and 99% prediction band) positive patient specimens. **B and C**). Included samples (n=106) with Ct values of the S, RdRP and N gene by the Allplex™ SARS-CoV-2/FluA/FluB/RSV assay with the corresponding results of whole genome sequencing or variant specific PCR. N501Y+, Del. H69/V70+ indicate presence of lineage B.1.1.7 **D.)** Nucleotide sequence alignment (MUSCLE) of N gene sequences from two exemplary patient samples (non-B.1.1.7 and B.1.1.7, respectively) and pCRII-TOPO_N-gene_D3 and _3L plasmids. Only the region containing the D3L mutation is shown (nucleotide position 1-12), with D3 highlighted in green (non-B.1.1.7) and 3L (B.1.1.7) highlighted in red. Translated amino acid sequences are shown above and below the nucleotide sequences. N gene sequences from the patient samples were extracted from whole-genome sequences.

**A)**

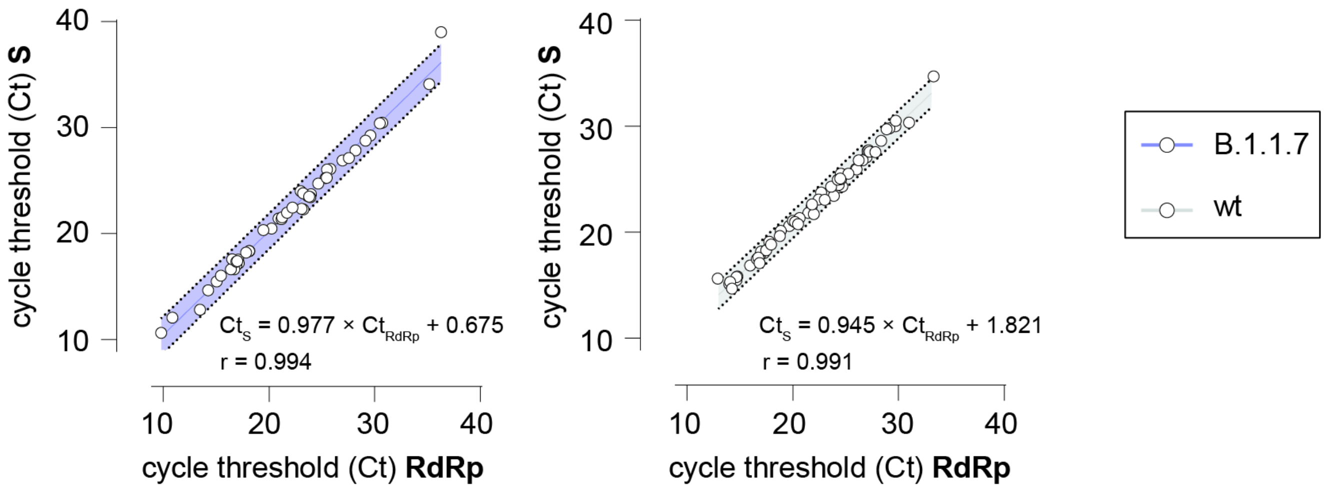

cycle threshold (Ct) S
40
30
20
10
$Ct_S = 0.977 \times Ct_{RdRp} + 0.675$
r = 0.994
10
20
30
40
cycle threshold (Ct) RdRp
$Ct_S = 0.945 \times Ct_{RdRp} + 1.821$
r = 0.991
B.1.1.7
wt

| B) | | Allplex™ SARS-CoV-2/FluA/FluB/RSV | | | Δ N/RdRP | Δ N/S | Lineage / variant PCR |
|---|---|---|---|---|---|---|---|
| | Sample | S | RdRP | N | | | |
| | patient 1 | 26.10 | 25.80 | 32.80 | **7.00** | **6.70** | B.1.1.7 |
| | patient 2 | 17.54 | 17.09 | 24.31 | **7.22** | **6.77** | B.1.1.7 |
| | patient 3 | 26.04 | 25.48 | 33.16 | **7.68** | **7.12** | B.1.1.7 |
| | patient 4 | 22.27 | 23.26 | 31.47 | **8.21** | **9.20** | B.1.1.7 |
| | patient 5 | 18.33 | 18.13 | 25.44 | **7.31** | **7.11** | B.1.1.7 |
| | patient 6 | 26.90 | 26.96 | 37.23 | **10.27** | **10.33** | B.1.1.7 |
| | patient 7 | 21.42 | 20.91 | 27.75 | **6.84** | **6.33** | B.1.1.7 |
| | patient 8 | 18.35 | 18.10 | 24.55 | **6.45** | **6.20** | B.1.1.7 |
| | patient 9 | 22.32 | 23.07 | 30.69 | **7.62** | **8.37** | B.1.1.7 |
| | patient 10 | 12.79 | 13.47 | 21.56 | **8.09** | **8.77** | B.1.1.7 |
| | patient 11 | 15.48 | 15.05 | 24.39 | **9.34** | **8.91** | B.1.1.7 |
| | patient 12 | 17.19 | 17.15 | 24.85 | **7.70** | **7.66** | B.1.1.7 |
| | patient 13 | 29.25 | 29.59 | - | **-** | **-** | N501Y+, Del. H69/V70+ |
| | patient 14 | 27.83 | 28.19 | 37.48 | **9.29** | **9.65** | B.1.1.7 |
| | patient 15 | 17.03 | 16.90 | 25.92 | **9.02** | **8.89** | B.1.1.7 |
| | patient 16 | 25.23 | 25.36 | 33.51 | **8.15** | **8.28** | B.1.1.7 |
| | patient 17 | 21.35 | 21.19 | 28.51 | **7.32** | **7.16** | N501Y+, Del. H69/V70+ |
| | patient 18 | 27.14 | 27.55 | 35.38 | **7.83** | **8.24** | N501Y+, Del. H69/V70+ |
| | patient 19 | 23.53 | 23.66 | 30.65 | **6.99** | **7.12** | B.1.1.7 |
| | patient 20 | 14.63 | 14.23 | 22.40 | **8.17** | **7.77** | B.1.1.7 |
| | patient 21 | 20.46 | 20.25 | 28.61 | **8.36** | **8.15** | B.1.1.7 |
| | patient 22 | 10.63 | 9.80 | 19.90 | **10.10** | **9.27** | B.1.1.7 |
| | patient 23 | 24.71 | 24.65 | 31.86 | **7.21** | **7.15** | B.1.1.7 |
| | patient 24 | 28.75 | 29.14 | - | **-** | **-** | B.1.1.7 |
| | patient 25 | 23.70 | 23.97 | 31.78 | **7.81** | **8.08** | B.1.1.7 |
| | patient 26 | 16.61 | 16.71 | 24.28 | **7.57** | **7.67** | N501Y+, Del. H69/V70+ |
| | patient 27 | 18.20 | 17.83 | 25.86 | **8.03** | **7.66** | N501Y+, Del. H69/V70+ |
| | patient 28 | 21.54 | 21.23 | 29.46 | **8.23** | **7.92** | N501Y+, Del. H69/V70+ |
| | patient 29 | 16.63 | 16.39 | 25.55 | **9.16** | **8.92** | N501Y+, Del. H69/V70+ |
| | patient 30 | 24.00 | 23.00 | 28.88 | **5.88** | **4.88** | N501Y+, Del. H69/V70+ |
| | patient 31 | 20.50 | 20.32 | 27.67 | **7.35** | **7.17** | N501Y+, Del. H69/V70+ |
| | patient 32 | 12.07 | 10.85 | 19.89 | **9.04** | **7.82** | N501Y+, Del. H69/V70+ |
| | patient 33 | 20.33 | 19.46 | 28.30 | **8.84** | **7.97** | N501Y+, Del. H69/V70+ |
| | patient 34 | 17.57 | 16.50 | 25.29 | **8.79** | **7.72** | N501Y+, Del. H69/V70+ |
| | patient 35 | 23.81 | 23.23 | 31.61 | **8.38** | **7.80** | N501Y+, Del. H69/V70+ |
| | patient 36 | 18.43 | 18.04 | 26.31 | **8.27** | **7.88** | B.1.1.7 |
| | patient 37 | 30.45 | 30.70 | - | **-** | **-** | N501Y+, Del. H69/V70+ |
| | patient 38 | 30.40 | 30.48 | - | **-** | **-** | N501Y+, Del. H69/V70+ |
| | patient 39 | 34.10 | 35.15 | - | **-** | **-** | N501Y+, Del. H69/V70+ |
| | patient 40 | 16.02 | 15.45 | **23.73** | **8.28** | **7.71** | B.1.1.7 |
| | patient 41 | 21.97 | 21.69 | **28.61** | **6.92** | **6.64** | B.1.1.7 |
| | patient 42 | 17.35 | 16.91 | **23.66** | **6.75** | **6.31** | B.1.1.7 |
| | patient 43 | 23.42 | 23.92 | **30.73** | **6.81** | **7.31** | B.1.1.7 |
| | patient 44 | 23.49 | 23.79 | **31.06** | **7.27** | **7.57** | B.1.1.7 |
| | patient 45 | 39.01 | 36.29 | - | **-** | **-** | B.1.1.7 |
| | patient 46 | 17.41 | 16.98 | **25.59** | **8.61** | **8.18** | B.1.1.7 |
| | patient 47 | 22.46 | 22.23 | **28.52** | **6.29** | **6.06** | B.1.1.7 |
| | patient 48 | 25.25 | 25.47 | **32.25** | **6.78** | **7.00** | B.1.1.7 |

| C) | | Allplex™ SARS-CoV-2/FluA/FluB/RSV | | | Δ N/RdRP | Δ N/S | Lineage / variant PCR |
|---|---|---|---|---|---|---|---|
| | Sample | S | RdRP | N | | | |
| | patient 49 | 15.81 | 14.76 | 13.62 | 1.14 | 2.19 | B.1.1.317 |
| | patient 50 | 16.85 | 15.97 | 14.79 | 1.18 | 2.06 | B.1.1.317 |
| | patient 51 | 30.01 | 29.72 | 27.62 | 2.10 | 2.39 | N501Y+, E484K+ |
| | patient 52 | 18.04 | 17.28 | 16.94 | 0.34 | 1.10 | N501Y-, Del. H69/V70- |
| | patient 53 | 24.24 | 24.37 | 23.87 | 0.50 | 0.37 | N501Y-, Del. H69/V70- |
| | patient 54 | 24.32 | 24.68 | 23.87 | 0.81 | 0.45 | N501Y-, Del. H69/V70- |
| | patient 55 | 15.46 | 14.67 | 14.47 | 0.20 | 0.99 | N501Y-, Del. H69/V70- |
| | patient 56 | 15.77 | 14.67 | 14.33 | 0.34 | 1.44 | N501Y-, Del. H69/V70- |
| | patient 57 | 17.83 | 16.91 | 15.00 | 1.91 | 2.83 | N501Y-, Del. H69/V70- |
| | patient 58 | 24.45 | 24.34 | 22.52 | 1.82 | 1.93 | N501Y-, Del. H69/V70- |
| | patient 59 | 17.44 | 16.59 | 15.95 | 0.64 | 1.49 | N501Y-, Del. H69/V70- |
| | patient 60 | 25.98 | 25.97 | 25.22 | 0.75 | 0.76 | N501Y-, Del. H69/V70- |
| | patient 61 | 29.81 | 29.14 | 27.36 | 1.78 | 2.45 | N501Y-, Del. H69/V70- |
| | patient 62 | 27.07 | 26.99 | 25.94 | 1.05 | 1.13 | N501Y-, Del. H69/V70- |
| | patient 63 | 28.62 | 28.35 | 26.22 | 2.13 | 2.40 | N501Y-, Del. H69/V70- |
| | patient 64 | 18.17 | 16.99 | 16.11 | 0.88 | 2.06 | N501Y-, Del. H69/V70- |
| | patient 65 | 20.57 | 19.65 | 18.50 | 1.15 | 2.07 | N501Y-, Del. H69/V70- |
| | patient 66 | 15.61 | 12.88 | 12.27 | 0.61 | 3.34 | N501Y-, Del. H69/V70- |
| | patient 67 | 22.45 | 21.80 | 21.14 | 0.66 | 1.31 | N501Y-, Del. H69/V70- |
| | patient 68 | 23.74 | 22.68 | 21.35 | 1.33 | 2.39 | N501Y-, Del. H69/V70- |
| | patient 69 | 21.06 | 20.01 | 18.79 | 1.22 | 2.27 | N501Y-, Del. H69/V70- |
| | patient 70 | 27.68 | 27.07 | 26.11 | 0.96 | 1.57 | N501Y-, Del. H69/V70- |
| | patient 71 | 20.12 | 18.87 | 17.69 | 1.18 | 2.43 | N501Y-, Del. H69/V70- |
| | patient 72 | 15.09 | 13.98 | 13.52 | 0.46 | 1.57 | N501Y-, Del. H69/V70- |
| | patient 73 | 25.91 | 26.03 | 23.74 | 2.29 | 2.17 | N501Y-, Del. H69/V70- |
| | patient 74 | 19.64 | 18.76 | 18.10 | 0.66 | 1.54 | N501Y-, Del. H69/V70- |
| | patient 75 | 18.28 | 17.49 | 16.77 | 0.72 | 1.51 | N501Y-, Del. H69/V70- |
| | patient 76 | 17.59 | 16.73 | 16.09 | 0.64 | 1.50 | N501Y-, Del. H69/V70- |
| | patient 77 | 19.08 | 17.77 | 16.12 | 1.65 | 2.96 | N501Y-, Del. H69/V70- |
| | patient 78 | 23.09 | 22.45 | 21.24 | 1.21 | 1.85 | N501Y-, Del. H69/V70- |
| | patient 79 | 18.83 | 17.95 | 16.56 | 1.39 | 2.27 | N501Y-, Del. H69/V70- |
| | patient 80 | 27.74 | 27.18 | 26.60 | 0.58 | 1.14 | N501Y-, Del. H69/V70- |
| | patient 81 | 15.29 | 14.08 | 13.34 | 0.74 | 1.95 | N501Y-, Del. H69/V70- |
| | patient 82 | 14.68 | 14.24 | 12.99 | 1.25 | 1.69 | N501Y-, Del. H69/V70- |
| | patient 83 | 26.78 | 26.60 | 25.85 | 0.75 | 0.93 | N501Y-, Del. H69/V70- |
| | patient 84 | 17.07 | 16.86 | 16.25 | 0.61 | 0.82 | N501Y-, Del. H69/V70- |
| | patient 85 | 25.51 | 24.55 | 23.89 | 0.66 | 1.62 | N501Y-, Del. H69/V70- |
| | patient 86 | 25.01 | 24.58 | 23.20 | 1.38 | 1.81 | N501Y-, Del. H69/V70- |
| | patient 87 | 23.42 | 23.88 | 22.09 | 1.79 | 1.33 | N501Y-, Del. H69/V70- |
| | patient 88 | 26.78 | 26.24 | 25.44 | 0.80 | 1.34 | N501Y-, Del. H69/V70- |
| | patient 89 | 27.59 | 27.28 | 26.16 | 1.12 | 1.43 | N501Y-, Del. H69/V70- |
| | patient 90 | 16.85 | 15.97 | 14.76 | 1.21 | 2.09 | N501Y-, Del. H69/V70- |
| | patient 91 | 34.70 | 33.32 | 31.63 | 1.69 | 3.07 | N501Y-, Del. H69/V70- |
| | patient 92 | 21.83 | 21.48 | 19.50 | 1.98 | 2.33 | N501Y-, Del. H69/V70- |
| | patient 93 | 24.28 | 23.63 | 22.82 | 0.81 | 1.46 | N501Y-, Del. H69/V70- |
| | patient 94 | 21.34 | 20.65 | 19.76 | 0.89 | 1.58 | N501Y-, Del. H69/V70- |
| | patient 95 | 23.06 | 23.01 | 21.90 | 1.11 | 1.16 | N501Y-, Del. H69/V70- |
| | patient 96 | 27.57 | 27.82 | 26.53 | 1.29 | 1.04 | N501Y-, Del. H69/V70- |
| | patient 97 | 21.71 | 21.98 | 20.94 | 1.04 | 0.77 | N501Y-, Del. H69/V70- |
| | patient 98 | 29.71 | 28.87 | 27.83 | 1.04 | 1.88 | N501Y-, Del. H69/V70- |
| | patient 99 | 20.96 | 20.20 | 19.30 | 0.90 | 1.66 | N501Y-, Del. H69/V70- |
| | patient 100 | 24.94 | 24.25 | 23.36 | 0.89 | 1.58 | N501Y-, Del. H69/V70- |
| | patient 101 | 30.33 | 30.99 | 28.11 | 2.88 | 2.22 | N501Y-, Del. H69/V70- |
| | patient 102 | 30.52 | 29.73 | 28.49 | 1.24 | 2.03 | N501Y-, Del. H69/V70- |
| | patient 103 | 25.52 | 25.26 | 24.44 | 0.82 | 1.08 | N501Y-, Del. H69/V70- |
| | patient 104 | 22.63 | 21.81 | 19.75 | 2.06 | 2.88 | N501Y-, Del. H69/V70- |
| | patient 105 | 25.07 | 24.48 | 23.45 | 1.03 | 1.62 | N501Y-, E484K+ |
| | patient 106 | 20.74 | 20.49 | 20.39 | 0.1 | 0.35 | B.1.351 |

**D)**

| | 1 | | | 12 |
|---|---|---|---|---|
| | M | S | D | T |
| patient 49 (non-B.1.1.7) | ATG | TCT | GAT | AAT |
| pCRII-TOPO_N-gene_D3 | ATG | TCT | GAT | AAT |
| patient 1 (B.1.1.7) | ATG | TCT | CTA | AAT |
| pCRII-TOPO_N-gene_3L | ATG | TCT | CTA | AAT |
| | M | S | L | T |
